# Autoantibodies Predictive of Atherosclerosis Progression and Statin Response in Juvenile-Onset SLE: A Biomarker Discovery Study

**DOI:** 10.64898/2026.03.24.26349192

**Authors:** Junjie Peng, Pierre Dönnes, Thomas McDonnell, Stacy P. Ardoin, Laura E. Schanberg, Laura B. Lewandowski, Elizabeth C. Jury, George A. Robinson, Coziana Ciurtin, the APPLE trial investigators, Childhood Rheumatology Research Alliance (CARRA)

## Abstract

**Importance:** Cardiovascular disease (CVD) is a major cause of morbidity/mortality in juvenile-onset systemic lupus erythematosus (JSLE), yet no reliable tools exist to stratify CVD-risk.

**Objective:** To identify serum biomarkers associated with atherosclerosis progression and response to atorvastatin.

**Design/Setting:** We used data/samples from a sub-cohort of the APPLE trial (2009) which investigated atorvastatin vs. placebo to reduce atherosclerosis progression in JSLE, measured by change in carotid intima-media thickness (CIMT), and conducted a baseline autoantibody diagnostic-accuracy biomarker study.

**Participants/Exposure:** APPLE trial participants (randomized 1:1 to atorvastatin vs. placebo) with matched baseline serum samples and stratified based on 36-month CIMT progression were included in the analysis.

**Main Outcomes and Measures:** Baseline serum autoantibodies were profiled using a functional proteomic platform (Sengenics, N=94). Empirical Bayes moderated t-test and Receiver Operating Characteristic (ROC) based logistic regression were applied to identify autoantibody signatures predictive of high vs. low atherosclerosis progression.

**Results:** Ninety-four children and young people with JSLE (age mean [SD] =15.3 [2.4] years; 73 [78%] female, 8 [8.5%] Asian, 23 [24.5%] Black, 43 [45.7%] White, and 20 [21.3%] Other) were evaluated. Autoantibody levels against six novel autoantigens (STK24, RAD23B, HDAC4, STAT4, SEPTIN9, NFIA) classified high vs. low CIMT progression in the placebo arm (combined AUC 0.87, 95% CI −0.75 to 0.96). In the atorvastatin arm, autoantibodies to eight autoantigens (ABI1, ATP5B, CSNK2A2, NRIP3, PRKAR1A, PDK4, BATF, NUDT2), distinguished the statin responders vs. non-responders (combined AUC 0.96, 95% CI −0.88 to 1). An additional 27-autoantibody signature predicted response/partial response to atorvastatin (AUC 0.88, 95% CI – 0.76 to 0.97). Protein–protein interaction analysis identified endothelial disruption and lipid infiltration as key atherosclerosis mechanisms in atorvastatin non-responders. Combining the autoantibody prediction models with disease parameters and a metabolic signature did not increase model performance in either placebo (AUC 0.81, 95% CI – 0.68 to 0.94 vs. 0.87, 95% CI −0.75 to 0.96) or sttin arms (AUC 0.84, 95% CI −0.73 to 0.95 vs. 0.88, 95% CI −0.76 to 0.97).

**Conclusions and Relevance:** This study identified novel autoantibody signatures for atherosclerosis progression and statin response in JSLE, with potential utility for precision medicine approaches for CVD-risk management.

**Key Points:** *Question:* Can functional proteomic analyses identify autoantibody signatures predictive of atherosclerosis progression and response to statin treatment in children and young people with juvenile-onset systemic lupus erythematosus?

*Findings:* Using baseline samples from the APPLE trial (1:1 RCT of atorvastatin vs placebo), we identified novel autoantibody profiles that accurately distinguished individuals with high versus low carotid intima-media thickness progression over three years in both placebo (AUC 0.87, 95% CI-0.75 to 0.96) and atorvastatin groups (AUC 0.96, 95% CI-0.88 to 1).

*Meaning:* Autoantibody signatures show strong potential for early risk stratification and for identifying those most likely to benefit from statin therapy.

## Introduction

Systemic Lupus Erythematosus (SLE) is a chronic inflammatory condition that markedly increases the risk of premature atherosclerosis and cardiovascular disease (CVD)-related mortality^1^ ^2^.

SLE with onset before 18 years of age (juvenile-onset SLE [JSLE]) presents with a more severe phenotype and higher comorbidity burden, including CVD^3^. In JSLE, both traditional and disease-related factors—particularly chronic inflammation, endothelial dysfunction, immune-cell vascular infiltration, and disrupted lipid metabolism—drive accelerated atherosclerosis^3–6^.

Sub-clinical atherosclerosis, which can be detected with non-invasive imaging techniques, has been found in ~32% of children and young people (CYP) with JSLE^7^. In the UK JSLE cohort (N=413), 12 CVD-related events were reported (~3%, mostly strokes)^8^, far exceeding rates in age-matched general populations (≈1:1,000 for CVD events^9^; ≈1:10,000 for strokes^10^). A literature review of SLE cases under 35 years reported myocardial infarction at a mean age of 24L±L6.4 years (range 5–35), underscoring the early onset of atherosclerosis in SLE^11^.

As statins are used widely for CVD-risk management, the APPLE trial (Atherosclerosis Prevention in Pediatric Lupus Erythematosus), was conducted to assess the efficacy and safety of atorvastatin in preventing subclinical atherosclerosis progression in JSLE, using carotid intima media thickness (CIMT) as primary endpoint ^12^. Although the primary endpoint was not met, APPLE analyses identified multiple traditional and non-traditional CVD-risk factors linked to CIMT progression^12–15^ and showed that premature atherosclerosis advances rapidly in CYP with JSLE.

There are currently neither reliable CVD-risk stratification tools for use in JSLE in clinical practice^16^, nor consensus among clinicians regarding the use of currently available tests^17^. Previous research identified serum metabolomic biomarkers predictive of CVD-risk in healthy CYP^18,19^ and in CYP with JSLE^5,6^, but they are not implemented in routine care.

This study aimed to identify novel autoantibody biomarkers predictive of atherosclerosis progression and statin response in JSLE, using a functional proteomic platform and well-curated APPLE trial samples. We hypothesised that such biomarkers could help stratify CVD-risk and support more targeted CVD-risk management.

## Methods

This diagnostic accuracy study was designed and reported in accordance with STARD 2015 guidelines (attached separately).

### Ethical approval and consent

The Duke Clinical Research Institute (Durham, NC) was the clinical- and data-coordinating centre, providing oversight of the APPLE study conduct, management, and statistical analysis. The study was conducted at 21 Childhood Arthritis and Rheumatology Research Alliance (CARRA) sites in North America. Local institutional review board approval was obtained, and all patients or their guardians gave informed consent and assent following local guidelines. No individuals were identifiable. ClinicalTrials.gov Identifier: NCT00065806. The Principal Investigator was Prof. Laura E. Schanberg, Duke University.

### Participants

This diagnostic accuracy (biomarker) cohort study included a subset of CYP with JSLE enrolled in the APPLE trial selected based on serum sample availability at baseline and matched complete datasets collected over the duration of the APPLE trial (N=94, 49 in the atorvastatin arm). Details about the APPLE trial protocol^12^ and cohort stratification based on CIMT progression over 36 months^20^ are included in the **Supplement.** Race and ethnicity were self-reported and investigated in the APPLE trial and this diagnostic accuracy study because JSLE severity is influenced by race and ethnicity. The “Other” race category included individuals who did not self-report as Asian, Black, or White. We reported only biological sex as collected according to the APPLE trial protocol.

### Biomarker analysis

Baseline serum samples from APPLE study participants stratified by 36-month CIMT as previously published^20^, were analysed using the Sengenics i-Ome Discovery platform, which profiles over 1,800 immune-relevant autoantibodies with high antigen specificity. Further methodological details are provided in the **Supplement**.

Lipid metabolomic data from matched serum samples, generated and analysed in our previous study^20^ were also available and incorporated into the composite model for predicting atherosclerosis progression.

### Data/Statistical Analyses

Statistical analyses were performed in R, with full details provided in the **Supplement** and figure legends. Significance was set at PL<L.05. Empirical Bayes moderated t-tests, ROC-based logistic regression, and sPLS-DA were used to identify predictive autoantibody profiles. The Youden method defined optimal sensitivity–specificity thresholds, and autoantibodies with AUCL>L0.60 were considered signature markers. Lipid metabolomics, autoantibody profiles, and baseline clinical parameters were integrated into the sPLS-DA models for CVD-risk prediction.

Details about *Model validation*, *Protein-protein interaction (PPI) analysis*, *Functional annotations, Exploratory correlation analyses* and *Logistic regression with treatment–biomarker interaction terms* are included in the **Supplement (eMethods).**

**Data analysis pipeline** is detailed in **Figure 1**.

**Figure 1.**
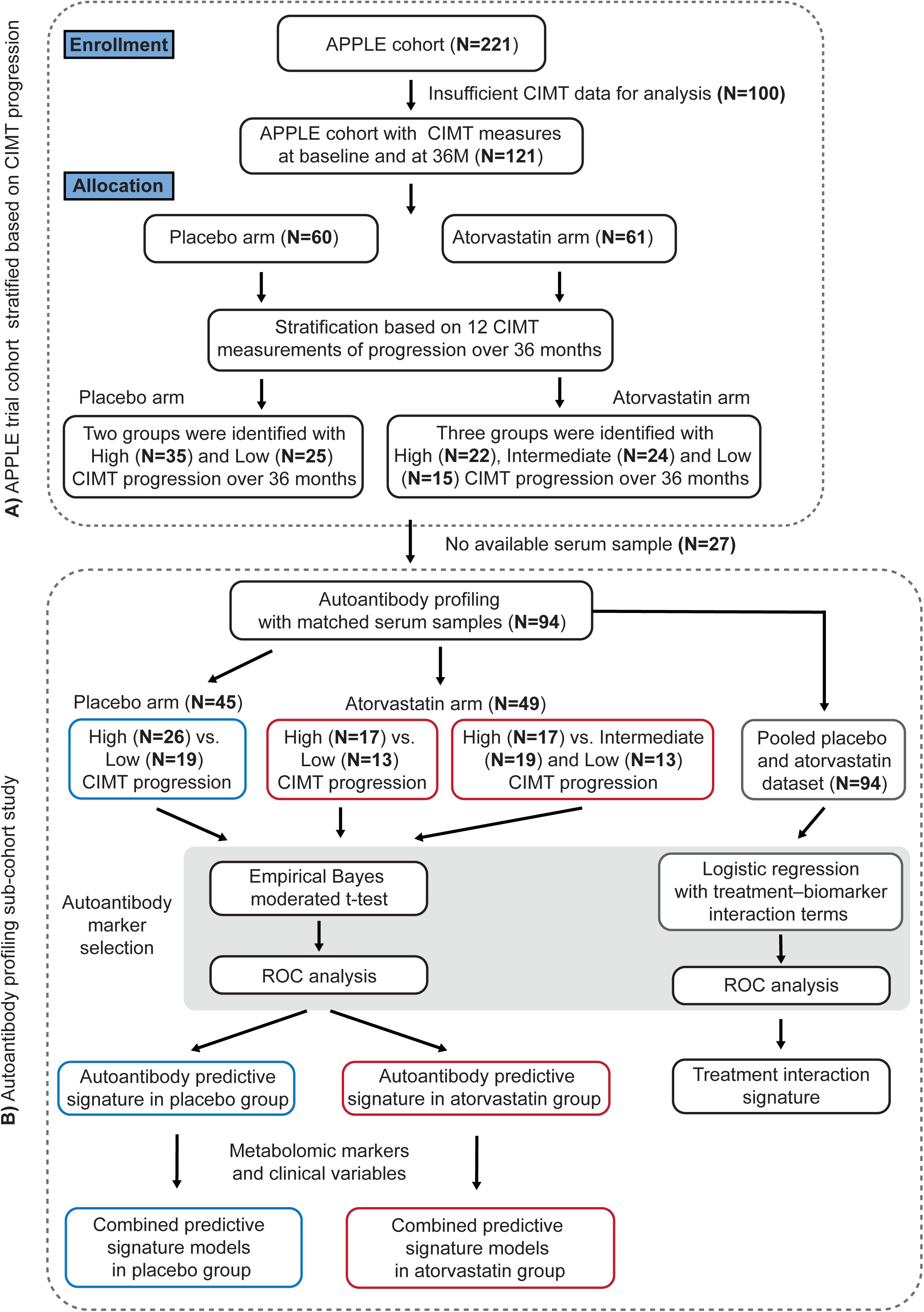
Study design and analysis plan flow diagram for the APPLE trial analyses. (A) The JSLE cohort was stratified based on CIMT progression over 36 months in both the placebo and atorvastatin arms^20^. (B) Summary of the pipeline analyses to define potential CVD-risk and treatment response functional proteomic biomarker signatures. *Abbreviations*: APPLE - Atherosclerosis Prevention in Pediatric Lupus Erythematosus clinical trial; CIMT - carotid intima-media thickness; CVD - cardiovascular disease; ROC - Receiver Operating Characteristic.

## Results

### Participant characteristics

Ninety-four participants recruited to the APPLE trial were included in this biomarker sub-cohort study. They were stratified based on CIMT progression over the duration of the APPLE trial using unsupervised hierarchical clustering analysis, as described in the **Supplement** and published previously^20^.

Baseline characteristics of the stratified APPLE trial sub-cohort analyzed in the current study (N=94) are shown in **Table 1**.

**Table 1.**
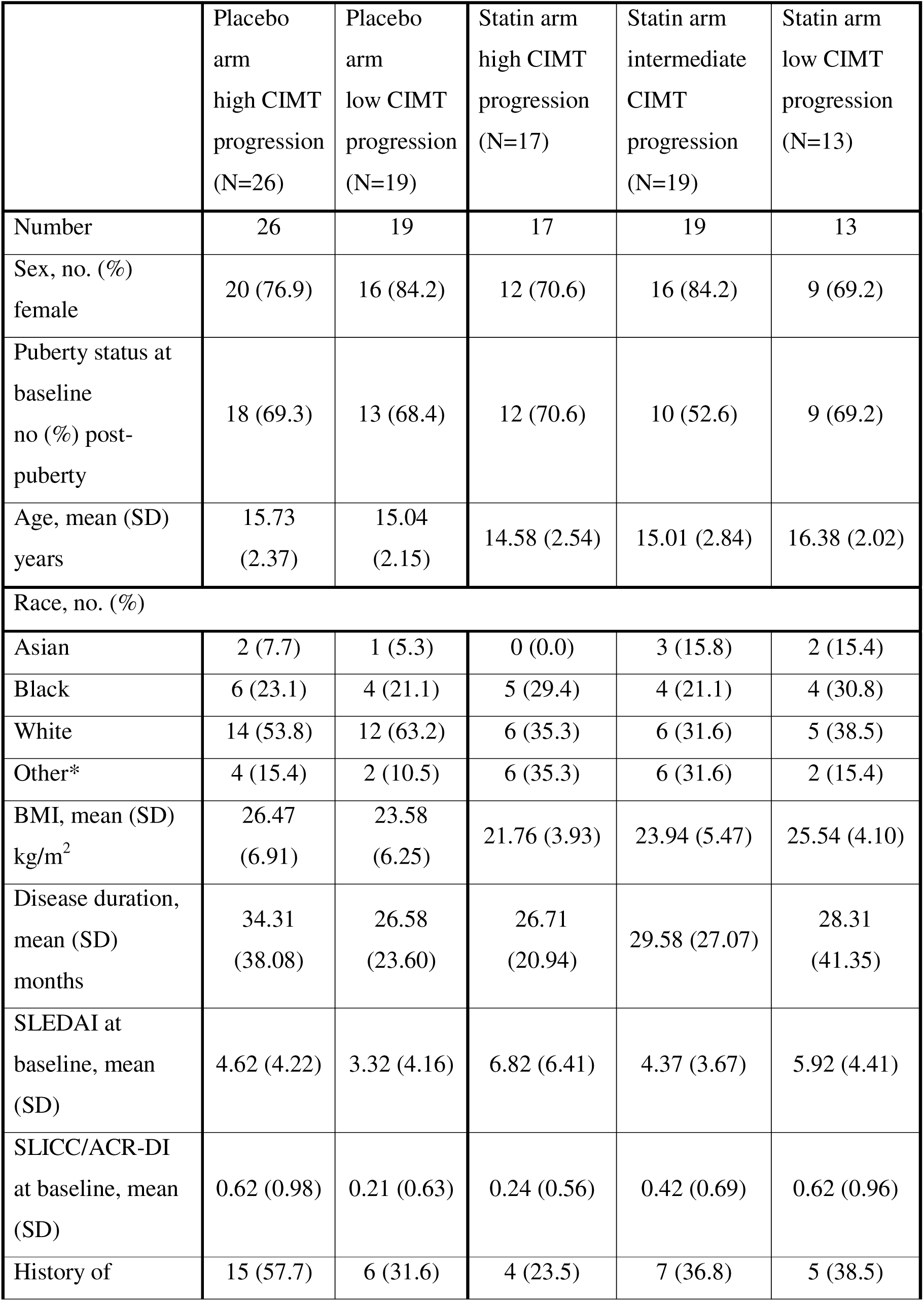
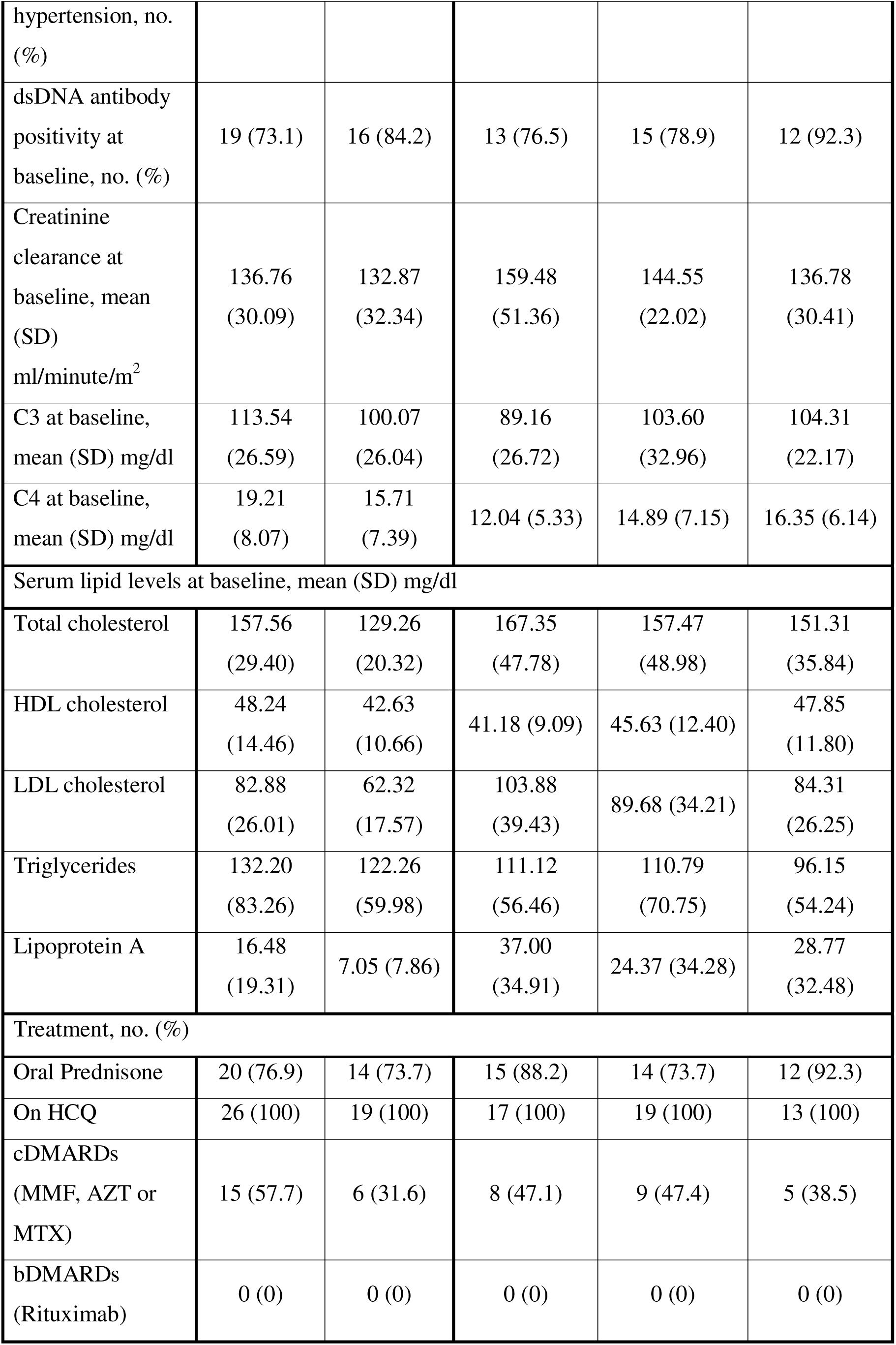
Demographic/clinical table of the APPLE trial sub-cohort (N=94) in the autoantibody profiling study. Abbreviations: BMI – Body mass index; SLEDAI – Systemic Lupus Erythematosus Disease Activity Index; SLICC/ACR-DI – Systemic Lupus International Collaborating Clinics/American College of Rheumatology Damage Index; dsDNA antibody – Anti-double-stranded ed DNA antibody; C3, C4 – complement fractions C3, C4; HDL – high-density lipoprotein; LDL – low-density lipoprotein; cDMARDs – conventional disease modifying antirheumatic drugs; bDMARDs – biologic; MMF – mycophenolate mofetil; AZT – azathioprine; MTX - methotrexate The “Other” race category included individuals who did not self-report as Asian, Black, or White.

### Distinct autoantibody signature predicted high vs. low CIMT progression in the placebo arm of the APPLE trial

Exploratory functional proteomic analysis identified six autoantibodies (P<.05; log2[fold change] >0.2) to be differentially expressed between CYP with high (N=26) vs. low (N=19) CIMT progression in the placebo arm of the APPLE trial. One autoantibody was significantly increased (STK24) and five decreased (RAD23B, HDAC4, STAT4, SEPTIN9, NFIA) (**Figure 2A-B**). Full names and functions of the autoantigens to which these novel autoantibodies were directed are listed in **eTable 1 in the Supplement.**

**Figure 2.**
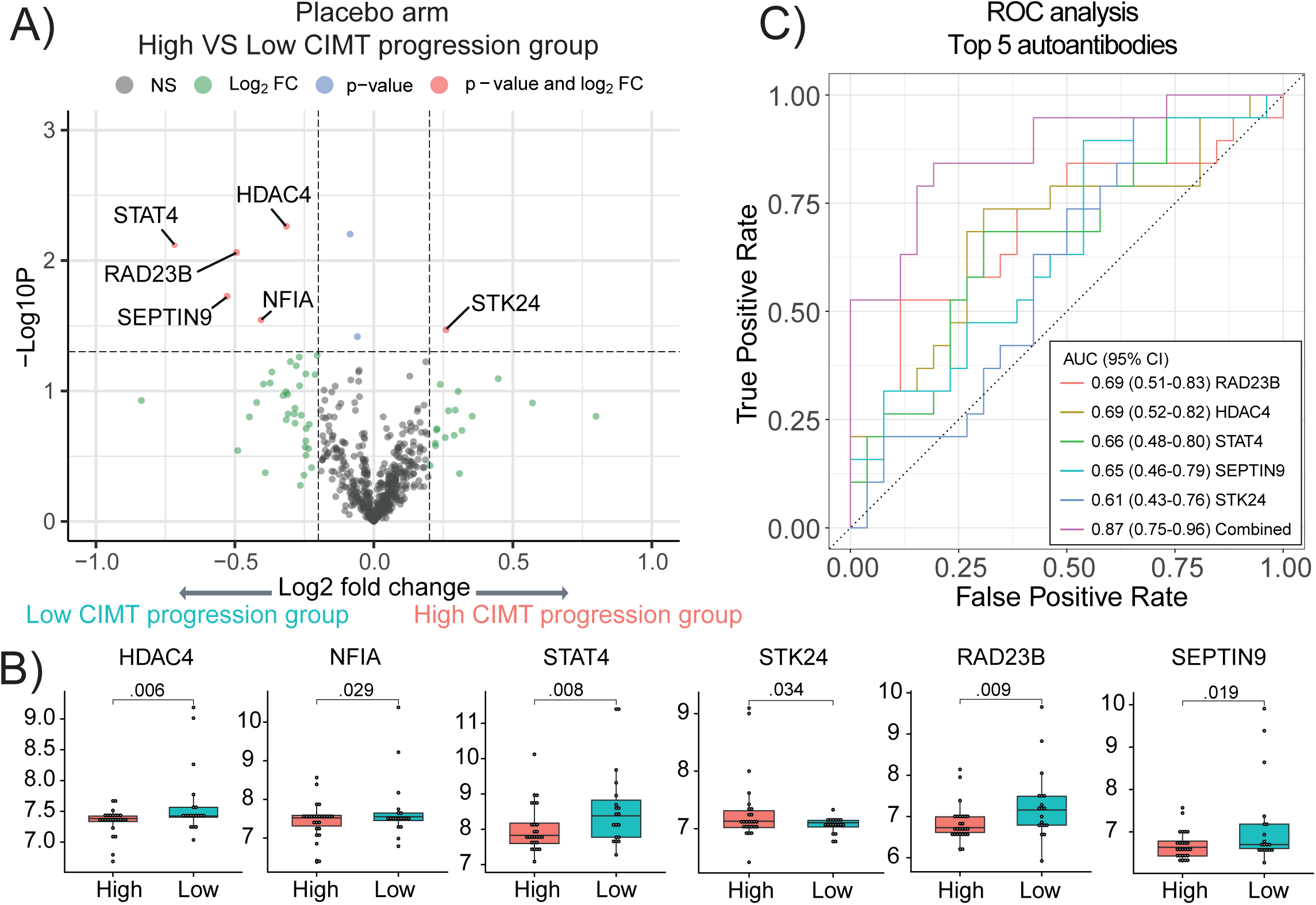
Baseline autoantibodies (N=579) compared across CIMT-progression groups in the placebo arm. A) Volcano plot displaying Log2 fold change of all autoantibodies and Log10 p values comparing high (N=26) and low (N=19) CIMT progression groups - placebo arm (P < .05; log2(fold change) > 0.2). Top five significant metabolites (AUC > 0.60) highlighted in red. B) Box and whisker plots showing the significant autoantibody levels of the high vs. low CIMT progression groups - placebo arm. Empirical Bayes moderated t-test. C) Individual and combined ROC analysis for discriminating high vs. low CIMT progression groups using the top five autoantibodies (individual AUC > 0.60). *Abbreviations*: AUC - area under the curve; CI - 95% confidence interval; CIMT- carotid intima-media thickness; ROC-Receiver Operating Characteristic.

Multivariable ROC analysis was performed to assess whether autoantibodies detected at baseline in CYP with JSLE could stratify patients according to their CIMT progression over the APPLE trial duration (high vs. low progression defined by unsupervised clustering analysis of CIMT patterns^20^ and **Supplement**). The top five autoantibodies (individual AUC cutoff >0.60, **eTable 2 in the Supplement)** demonstrated a combined AUC of 0.87 (95% CI-0.75 to 0.96) (**Figure 2C**), outperforming the accuracy of each autoantibody alone in predicting atherosclerosis progression in JSLE.

### Distinct autoantibody profile predicts response to statin treatment in the APPLE trial

The same analysis pipeline was employed to examine the baseline autoantibody profiles of CYP in the atorvastatin arm of the APPLE trial, comparing the high CIMT progression (statin non-responders, N=17) against low CIMT progression groups (statin responders, N=13) (Figure. 1). Eight autoantibodies were significantly differentially expressed (P< .05; log2[fold change] >0.2), with four upregulated (antibodies against ABI1, ATP5B, CSNK2A2, NRIP3) and four downregulated (antibodies against PRKAR1A, PDK4, BATF, NUDT2) in the high vs. low CIMT progression statin group (**Figure 3 A-B**). The full names and known functions of these autoantigens are listed in **eTable 3 in the Supplement**. ROC analysis in univariate logistic regression generated AUC values for the individual antibodies ranging from 0.47 to 0.74, while multivariable ROC analysis using the top five performing antibodies (individual AUCs >0.60) yielded a combined AUC of 0.96 (95% CI −0.88 to 1) (**eTable 4 in the Supplement** and **Figure 3C**).

**Figure 3.**
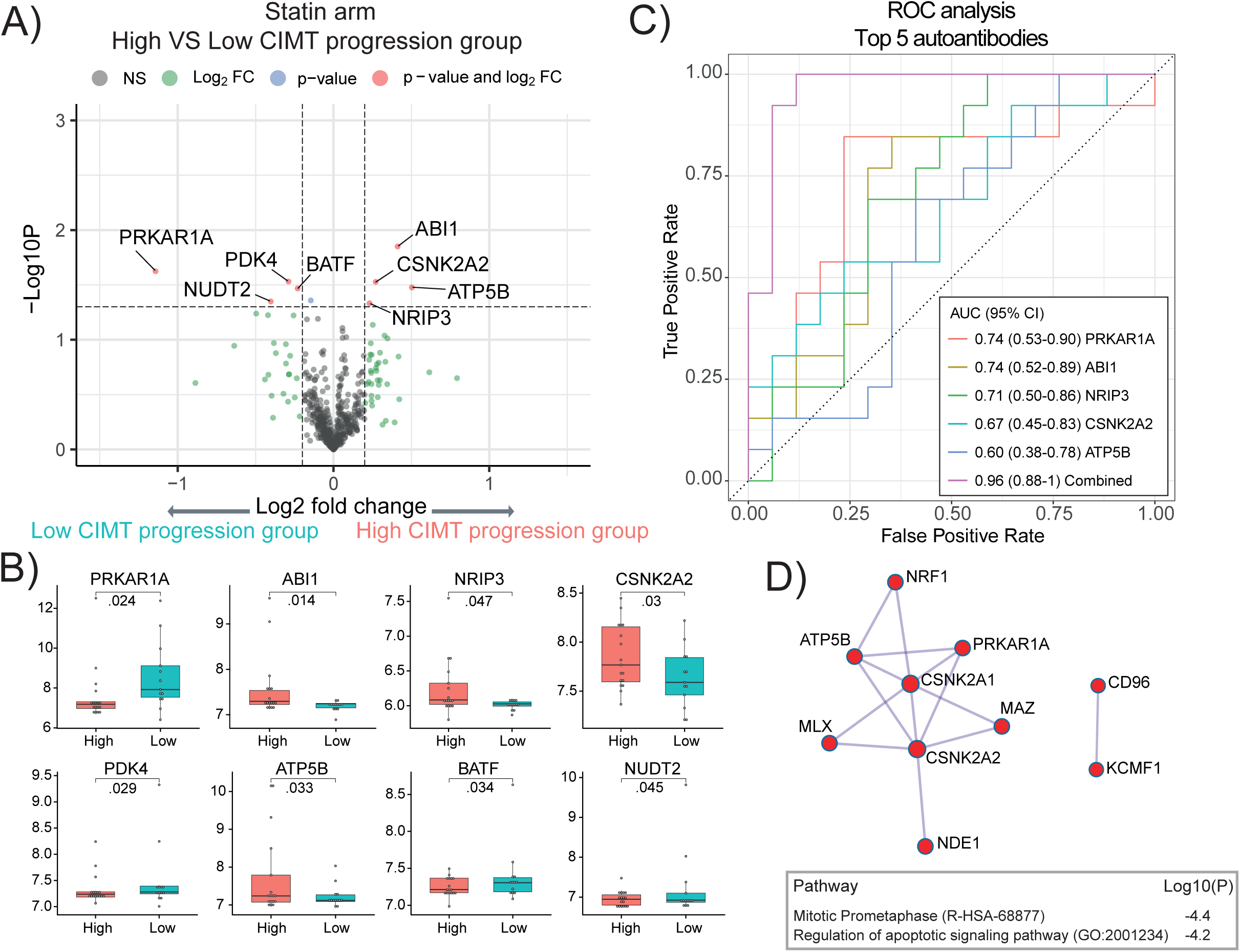
Statin-arm CIMT-progression groups compared by baseline autoantibody profiles (N=579). A) Volcano plot displaying Log2 fold change of all autoantibodies and Log10 p values comparing high (N=17) and low (N=13) CIMT progression groups - atorvastatin arm (P <.05; log2(fold change) > 0.2). B) Box and whisker plots showing the significantly distinct autoantibody levels in the high vs. low CIMT progression groups - atorvastatin arm. Empirical Bayes moderated t-test. C) Individual and combined ROC analyses for discriminating high vs. low CIMT progression groups using the top five autoantibodies (individual AUC > 0.60). D) Protein–protein interaction (PPI) network constructed using the 26 autoantibodies (excluding dsDNA) predictive of high CIMT progression in the statin arm of the APPLE trial. Enriched pathways identified from the PPI network are displayed in the box. *Abbreviations*: AUC - area under the curve; CI - 95% confidence interval; CIMT - carotid intima-media thickness; ROC - Receiver Operating Characteristic.

To increase statistical power and assess autoantibody patterns linked to atherosclerosis progression despite statin therapy, the low and intermediate CIMT-progression groups were combined into a moderate-progression group (N=32) (**Table 1**). This group was compared with high-progression non-responders, identifying 21 differentially expressed antibodies, 13 of which were upregulated (ABI1, CSNK2A2, MLX, FLI1, NRF1, ERG, SNRPA, dsDNA, RUNX1T1, CSNK2A1, MAZ, HOMER2, LIN28A) and 8 were downregulated (NAP1L3, CD96, KCMF1, EHF, CLK3, PPP1R2P9, NDE1, GFAP) in the high vs. moderate CIMT progression groups (individual AUCs 0.59 to 0.75) (**eFigure 1A-C,** and **eTables 3-4 in the Supplement**). Multivariable ROC analysis using the top five performing antibodies (individual AUC >0.69) yielded a combined AUC of 0.88 (95% CI −0.76 to 0.97) (**eFigure 1C in the Supplement**). As expected, combing the low and intermediate CIMT progression groups increased the cohort heterogeneity and led to reduced performance in the ROC analysis.

Predictive autoantibody signatures were then examined for links to atherosclerosis mechanisms. Due to the small placebo signature, PPI analysis was limited to the statin signature, revealing several interconnected autoantibody targets associated with CIMT progression despite treatment, including CSNK2A1 and CSNK2A2, PRKAR1A, ATP5B, MLX, NDE1, NRF1, MAZ, KCMF1, and CD96. Enriched pathway analysis indicated coordinated roles in processes such as mitotic prometaphase and regulation of apoptosis, which may contribute to endothelial junctional disruption through altered cell division and death (**Figure 3D**).

### Integration of multiomic signatures for prediction of atherosclerosis progression and atorvastatin response in JSLE

To assess the contribution of JSLE-related inflammation and dyslipidaemia to subclinical atherosclerosis, we evaluated whether perfectly matched baseline metabolomic profiles (previously published^20^) and autoantibody signatures from the same serum samples could identify CYP with high CIMT progression in the placebo arm. Overall, 88L% (23/26) of high-progressors had at least one of the six previously identified upregulated lipid metabolites^20^, and 100L% had at least one elevated autoantibody marker (**Figure 4A**). Only a small subset (N=3) showed the autoantibody signature alone, supporting the added value of combining both biomarker types for improved CVD-risk stratification.

**Figure 4.**
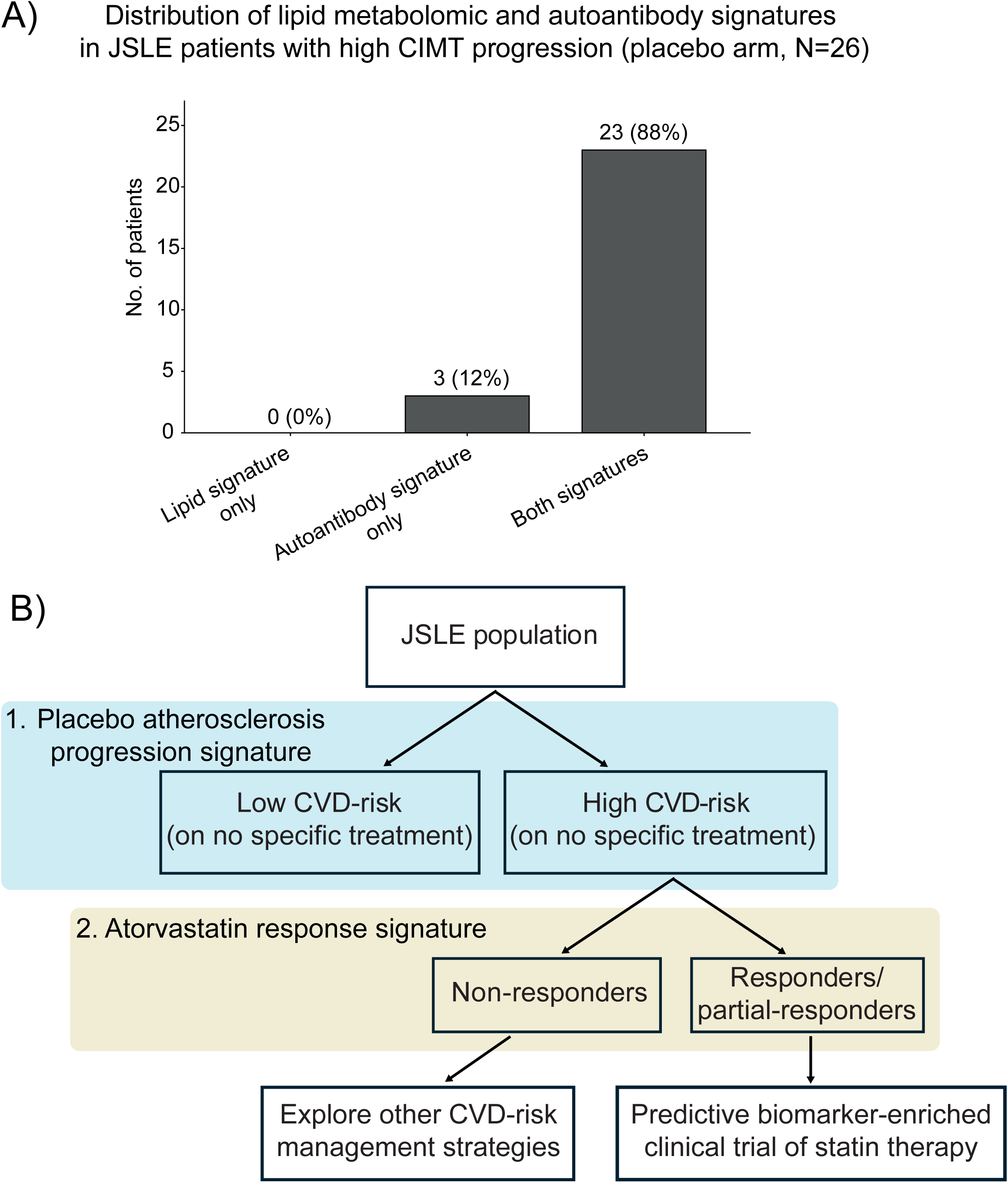
Distribution of serum-based predictive signatures among high-risk JSLE patients in the placebo arm and proposed two-step CVD-risk stratification. **A)** Distribution of lipid metabolomic and autoantibody signatures among JSLE patients with high carotid intima–media thickness (CIMT) progression in the placebo arm (N=26). Bars represent the number of patients positive for the lipid metabolomic signature only, the autoantibody signature only, or both signatures. The lipid metabolomic signature was defined by meeting at least one of the selected lipid metabolites (S-LDL-L, M-LDL-FC, S-LDL-C, S-LDL-PL, Total-C, Total-CE) above their published cut-offs^20^. The autoantibody signature was defined by positivity for at least one of the selected autoantibodies (RAD23B, HDAC4, STAT4, SEPTIN9, STK24, NFIA) above their cut-offs described in **eTable2**. **B)** Proposal of a two-step risk stratification and treatment strategy for atherosclerosis risk in JSLE. Flow chart shows the potential clinical implementation of a two-step strategy to first identify CYP with JSLE at risk of high atherosclerosis progression who can be targeted by any type of CVD-risk management strategy, and secondly, select with high accuracy the individuals with JSLE more likely to benefit from statin treatment as CVD-risk management strategy.

To develop clinically applicable predictive models for atherosclerosis progression and atorvastatin response, we integrated routinely collected clinical and laboratory data with matched baseline lipid-metabolomic profiles^20^ and autoantibody signatures from the APPLE cohort.

Integration analysis included the five top placebo signature antibodies (RAD23B, HDAC4, STAT4, SEPTIN9, STK24 **eTable 2 in the Supplement**), the top six lipid metabolites described previously^20^ (S-LDL-L, M-LDL-FC, S-LDL-C, S-LDL-PL, Total-C, Total-CE), and 21 JSLE variables measured at APPLE trial enrollment (age, sex, disease duration, body mass index (BMI), disease activity (SLEDAI) and damage (SLICC) scores, puberty status; serum LDL cholesterol, total cholesterol, high sensitivity C-Reactive Protein (hsCRP), ESR, and C3 and C4 (complement levels), as well as hemoglobin, white blood cell and platelet counts, and binary outcomes, dsDNA, anti-Ro and anti-La seropositivity, urine proteinuria and baseline treatment with glucocorticoids). The multivariable model successfully discriminated between CYP with high vs. low CIMT progression in the placebo arm (**eFigure 2A in the Supplement**), achieving an AUC of 0.81 (95% CI −0.68 to 0.94) under 5-fold cross-validation (**eFigure 2B in the Supplement**); however, this model did not outperform the autoantibody prediction model (AUC = 0.87, 95% CI −0.75 to 0.96, **Figure 2C**).

Similarly, in the atorvastatin arm, the integrated analysis included 25 antibodies with AUC >0.60 (**eTable 4 in the Supplement)** and the 21 JSLE variables (as above) (**eFigure 2C in the Supplement).** Since we did not identify a metabolic signature of statin response in the APPLE trial^20^, the analysis could not include lipid/metabolite biomarkers. The model differentiated individuals with high CIMT progression (statin non-responders) from those with intermediate or low progression (statin partial-responders and responders) with an AUC of 0.84 (95% CI-0.73 to 0.95) (**eFigure 2D in the Supplement**), highlighting no improved performance compared to the autoantibody signatures alone (AUC 0.88, 95% CI −0.76 to 0.97, **eFigure 1C in the Supplement**).

Finally, correlations between baseline serum autoantibodies and clinical parameters in both the placebo and atorvastatin groups were identified (**eFigure 3A–B in the Supplement**). In the placebo arm, no autoantibodies correlated with SLEDAI or SLICC scores. However, STK24 showed positive correlations with C4 levels and proteinuria; RAD23B correlated positively with ESR and negatively with anti-La antibodies; and STAT4 correlated negatively with disease duration. In contrast, in the atorvastatin arm, numerous baseline autoantibodies demonstrated significant associations with clinical measures. Notably, EHF and LIN28A correlated positively with SLEDAI, whereas CSNK2A1 and CLK3 correlated negatively. The robust performance of these internally validated integrated models, though not exceeding the autoantibody signatures, supports their use in clinical trial design to identify JSLE groups most likely to benefit from CVD-risk interventions or statin therapy, advancing personalised treatment strategies.

To examine whether baseline autoantibody levels predicted atherosclerosis progression differently across treatment groups, we used pooled logistic regression models incorporating treatment–biomarker interaction terms in the full cohort (atorvastatin and placebo arms combined). CIMT progression status was defined based on the CIMT changes over 36 months (**eFigure 4A in the Supplement**). Interaction analysis identified twelve baseline autoantibodies with nominally significant treatment–biomarker interaction effects (P < .05), yielding a combined AUC of 0.61 (95% CI-0.49 to 0.72) (**eFigure 4B-D in the Supplement).** Six of the twelve biomarkers were shared with those detected in the within-arm analyses, (**eTable 5 in the Supplement**), indicating partial overlap between the pooled and stratified models, although the pooled approach showed weaker overall performance.

## Discussion

This biomarker cohort study using data and samples from CYP with JSLE enrolled in the APPLE trial successfully identified a distinct novel autoantibody signature for high atherosclerosis progression in JSLE. Six key autoantibodies (against STK24, RAD23B, HDAC4, STAT4, SEPTIN9, NFIA) showed robust performance in multivariable ROC analysis, outperforming previously identified metabolomic markers of atherosclerosis progression using the same APPLE trial cohort^20^.

The six novel autoantibodies indicating atherosclerosis progression in JSLE, although not previously described in either SLE or JSLE, are key players in the cellular dynamics relevant for atherosclerosis development. Importantly, the corresponding autoantigens are involved in critical cellular processes, ranging from signal transduction and gene expression to DNA repair and cytoskeletal organization, in addition to lipid metabolism^21–25^, and potentially can be manipulated therapeutically. Therapeutic targeting of the JAK–STAT pathway has shown promise in SLE, with tofacitinib improving vascular function and lipid metabolism, and providing benefits in terms of immune modulation in individuals carrying the STAT4 risk allele^26^. HDAC4 and NFIA are identified as therapeutic targets for CVD, with existing interest in HDAC4 inhibitors^27^. CD40L blockade (e.g. dapirolizumab pegol) may lead to decreased STK24 expression and activity through their shared downstream NF-κB/IL-17 signalling^28^, while targeting STK24 may attenuate CD40L-driven NF-κB activation, potentially providing benefits for both SLE-related inflammation and atherosclerosis risk.

This autoantibody profiling analysis provided the first biomarkers predictive of statin benefit in slowing atherosclerosis progression in JSLE, despite the absence of a corresponding metabolomic signature^20^. Their known function, along with PPI enrichment analysis from the statin arm, suggest that lipid-independent pathways contribute to atherosclerosis progression in JSLE despite statin treatment. The eight novel autoantibodies defining the response signature of statins are also potentially relevant for lipid metabolism and atherosclerosis^29–38^. Notably, increased levels of autoantibodies to ABI1 and CSNK2A2 (CK2 component) emerged in our analysis as shared predictive markers of response/partial response to statin therapy. Both proteins can be therapeutically manipulated, with pan-NADPH oxidase (NOX) inhibitors (downstream of ABI1 signalling) found to reduce atherosclerotic plaque size, vascular inflammation, and oxidative stress^39^, and CK2 inhibitors being currently investigated as anti-inflammatory agents in atherosclerosis^40^. Additionally, ATP5B, an apolipoprotein A-I receptor with a role in liver lipid transport^41^, may also be mechanistically relevant as a statin-response biomarker, particularly given the availability of ATP5B inhibitors currently evaluated for other indications^42^.

Although we cannot determine whether these autoantigens directly drive atherosclerosis, the findings highlight broader pathways worth exploring in JSLE. Future work will clarify whether these autoantibodies are neutralizing, pro-atherosclerotic, or protective, and whether statins modify their activity. Notably, most placebo-signature autoantigens were linked to atherosclerosis-related processes, and their autoantibodies were reduced in high CIMT progressors, suggesting possible neutralizing roles.

In the placebo arm, only a few autoantibodies showed isolated associations with JSLE parameters. In contrast, in the atorvastatin arm, several baseline antibodies involved in transcriptional regulation (EHF, LIN28A) and kinase-mediated signalling (CSNK2A1, CLK3) correlated with disease activity. This indicates that the predictive value of SLEDAI for atherosclerosis progression depends on the underlying immunological milieu, with baseline disease activity relating to future CIMT progression only when accompanied by specific autoantibody patterns. Autoantibodies against RAD23B (placebo arm) and NDE1 and KCMF1 (atorvastatin arm) were downregulated in high-atherosclerosis progression groups, and their negative correlations with baseline cholesterol levels imply that reduced immune recognition of proteins involved in DNA repair, centrosome regulation, and ubiquitin-mediated turnover may reflect metabolic or inflammatory states characterised by lower lipid levels and altered cellular stress responses.

Notably, the addition of JSLE-related biomarkers and metabolomic signature did not improve the performance of the serum autoantibody predictive models for atherosclerosis progression in either the placebo arm (AUC 0.81, 95% CI-0.68 to 0.94 vs 0.87, 95% CI-0.75 to 0.96) or in the statin arm (AUC 0.84, 95% CI-0.73 to 0.95 vs. 0.88, 95% CI-0.76 to 0.97 for the combined responder and partial responder signatures). This highlights JSLE heterogeneity and the inability of routinely collected parameters to predict with accuracy high CVD-risk in CYP with JSLE or predict response to statins.

Although chronic JSLE therapies may influence the immune–metabolic milieu relevant to atherosclerosis progression, baseline treatment use was balanced across trial arms, ensuring any such effects were evenly distributed. Baseline autoantibody signatures therefore reflect predictors of CIMT progression and statin response rather than treatment effects, an inherent strength of the randomized design. Consistent with this, our previous work (in a larger sample^20^) showed no treatment-related differences in atherosclerosis progression in either arm. Disease activity and damage accrual also did not differ across CIMT progression groups, indicating that CIMT progression in this cohort is independent of disease severity or damage burden and does not account for the baseline biomarker associations observed.

Although the APPLE trial was completed 17 years ago, its findings remain directly relevant because no meaningful therapeutic shift has occurred in JSLE during that period. Belimumab is the only biologic licensed for JSLE, and it was not approved until 2019 (FDA) and 2021 (EMA). Consequently, for the entire post-APPLE era, most CYP with JSLE have continued to receive the same treatments as APPLE participants. No other biologics or novel drug classes are approved for this population, and real-world belimumab use in children appears very modest; the only paediatric belimumab trial (PLUTO) enrolled just 93 children worldwide^43^, underscoring how small the eligible population is. As a result, the treatment exposures shaping cardiovascular risk today remain broadly aligned with those in the APPLE trial.

Analyses conducted separately within each treatment arm identified prognostic biomarkers associated with 3-year CIMT progression under either statin or placebo. These models perform well (AUC 0.87, 95% CI-0.75 to 0.96 for placebo and AUC 0.96, 95% CI-0.88 to 1 for atorvastatin) because they estimate the link between baseline biology and outcome within a stable treatment environment. In contrast, the pooled interaction model estimates whether baseline biomarkers modify the difference in CIMT progression between statin and placebo. Because statin therapy alters vascular biology over time^44^, baseline biomarkers may only partially capture the pathways that drive onLtreatment CIMT change. This asymmetry reduces predictive performance and increases uncertainty in interaction estimates (AUC 0.61, 95% CI-0.49 to 0.72). Additionally, among the autoantibodies identified, only a minority targeted proteins with any plausible relevance to atherosclerosis. PHLDA1, PTGDR and, to a lesser extent, IPMK map to pathways involved in vascular inflammation, prostaglandin signalling, or immune–metabolic regulation^45,46^, providing some biological rationale for their association with CIMT progression. In contrast, GAGE2B/2C, PABPC1 and PHKG2 have no established mechanistic links to atherosclerosis or vascular biology. For these reasons, interaction analyses in randomized clinical trials are typically exploratory and hypothesisLgenerating rather than definitive tools for treatment selection. Longitudinal biomarker measurements and dynamic models of treatment response would be required to further characterize these relationships.

Our study is limited by the modest sample size available from the APPLE cohort. The autoantibody associations observed were internally consistent, biologically plausible, and present at baseline before treatment allocation, providing the first candidate biomarkers identified in a rigorously phenotyped, randomized JSLE cohort. Mechanistic interpretation and longitudinal effects of disease activity, immunosuppression, or statin therapy were beyond scope. Because strict FDR correction eliminated all signals and increased typeLII error, nominal PL<L.05 thresholds were used to highlight exploratory candidates linked to CIMT progression and treatment response. Several of these autoantibodies map to pathways relevant to vascular inflammation, immune regulation, or metabolism. These findings are hypothesis-generating and require validation in larger independent cohorts.

### Proposal of a two-step strategy for CVD-risk stratification and therapeutic decisions in JSLE

Our findings need validation in larger studies, but they support a two-step CVD-risk stratification approach in JSLE (**Figure 4B**). If confirmed, JSLE cohorts could first be stratified by predicted natural atherosclerosis progression using a validated autoantibody signature alone or the integrated serum lipid and autoantibody signature model identified in the placebo arm of the APPLE trial. We propose that CYP at low atherosclerosis risk could be monitored annually, while those at high risk could be further stratified using the second predictive model from the atorvastatin arm, based on autoantibody signatures of statin response, partial response, or the integrated signature. This approach could support tailored therapy—avoiding overtreatment in low-risk CYP and identifying those most likely to benefit from statins—while directing high-risk, likely non-responders toward alternative interventions. These predictive autoantibody models could support biomarker-enriched cohort selection for future interventional trials, though further validation is needed. This work may help close a key clinical gap by informing evidence-based strategies to improve CVD-risk management and address the top cause of mortality in JSLE in developed countries.

## Conclusions

This study found two distinct serum biomarker signatures comprising novel autoantibodies that showed high performance distinguishing individuals with JSLE at high risk for atherosclerosis progression as well as responders to statin treatment. Supported by robust biomarker performance and standardized CIMT assessment within the APPLE trial, these findings point to promising avenues for clinical application and could help address a key unmet need by improving identification of high-risk CYP and informing selection of those most likely to benefit from statins, while also guiding future research into targeted approaches for JSLE-associated atherosclerosis.

## Supporting information

Non-author collaborators

## Acknowledgements

We would like to thank all participants and hospital sites that recruited patients for the APPLE study. We would like to thank CARRA for support of this project and review of the manuscript.

## Collaborators

Excel file attached

## Access to data and data analysis

JP and CC had full access to all the data in the study and take responsibility for the integrity of the data and the accuracy of the data analysis.

## Data sharing statement

The APPLE clinical trial study protocol and results are publicly available. The study has been reported according to the **Consolidated Standards of Reporting Trials (CONSORT)** reporting guidelines (PMID: **22031171**). Metabolomic analyses of the APPLE trial are also publicly available (PMID **37786302**). Data used for all the complementary analyses included in this manuscript and the analytic codes will be available upon reasonable request after February 2026 (as data are protected by a patent application).

