## Supplementary material for "Autoantibodies Predictive of Atherosclerosis Progression and Statin Response in Juvenile-Onset SLE: A Biomarker Discovery Study"

**Supplement**

**Novel Autoantibodies Predictive of Atherosclerosis Progression and Statin Response in Juvenile-Onset Systemic Lupus Erythematosus – a Biomarker Discovery Study**

### Junjie Peng, PhD ^1,2^, Pierre Dönnes, PhD^3^, Thomas McDonnell, PhD^1^, Stacy P. Ardoin, MD^4^, Laura E. Schanberg, MD^5^, Laura Lewandowski, MD^6^, Elizabeth C. Jury, PhD^1^ †, George Robinson, PhD^1,2^ †, Coziana Ciurtin, PhD^1,2,7^ †, for the APPLE trial investigators and Childhood Rheumatology Research Alliance (CARRA)

† Share senior authorship

Number and title of each element included in the document:

1. **eMethods**
2. **eFigure 1.** Baseline serum autoantibodies (N=579 after data cleaning) comparisons between distinct CIMT progression groups (high vs. low and intermediate, identifying non-responders vs. responders and partial responders to statin) - atorvastatin arm.
3. **eFigure 2.** Sparse Partial Least Squares Discriminant Analysis (sPLS-DA) using identified autoantibody signature, identified lipid metabolomic signature and clinical variables to predict CVD-risk in placebo and statin arms.
4. **eFigure 3.** Chord plot showing exploratory correlations between baseline autoantibody signature and clinical features in the APPLE trial sub-cohort (N=94).
5. **eFigure 4.** Treatment–biomarker interaction logistic regression analysis in the APPLE trial sub-cohort.
6. **eTable 1** Names and functions of the top significantly differentially expressed autoantibodies identified in the high vs. low CIMT progression groups in the placebo arm.
7. **eTable 2.** Performance of the top significantly differentially expressed autoantibodies in stratifying atherosclerosis progression risk in the placebo arm.
8. **eTable 3.** Names and functions of the top significantly differentially expressed autoantibodies in stratifying atherosclerosis progression risk in the atorvastatin arm (includes both comparisons between the high vs. low CIMT progression groups (non-responders vs. responders to statins) and the combined high vs. low/moderate progression groups (non-responders vs. responders and partial responders to statin).
9. **eTable 4.** Performance of the top significantly differentially expressed autoantibodies in stratifying atherosclerosis progression risk in the atorvastatin arm (includes both comparisons between the high vs. low CIMT progression groups (non-responders vs. responders to statins) and the combined high vs. low/moderate progression groups (non-responders vs. responders and partial responders to statin).
10. **eTable 5.** Names and functions of the top autoantibodies (P < 0.05) identified in treatment–biomarker interaction logistic regression analysis.

**eMethods:**

***Participants:***

The APPLE trial enrolled 221 CYP with JSLE (age 10-18 at inclusion) who met the ACR 1997 revised classification criteria, recruited from 21 sites in North America, and followed for 36 months. Subjects were randomized 1:1 to receive either placebo (N=108) or atorvastatin (N=113), upon informed consent/assent as age and developmentally appropriate. All subjects met well defined inclusion/exclusion criteria as per published protocol^1^.

A total of 121 CYP with JSLE recruited to the APPLE trial who had available matched baseline serum samples and complete clinical, serological, and vascular imaging data were included in this biomarker discovery study. Access to these data and samples from the JSLE cohort was facilitated by an international collaboration with CARRA and APPLE trial investigators (USA).

CYP with JSLE have been stratified based on CIMT progression over 36 months in both the placebo and atorvastatin arms, using unsupervised hierarchical clustering, as we published before. This analysis stratified the CYP with JSLE recruited to the placebo-arm into two distinct groups with high (N=35) and low (N=25) CIMT progression over 36 months, and the CYP enrolled into the atorvastatin-arm into three distinct groups with high (N=22), intermediate (N=24) and low (N=15) CIMT progression over 36 months, as we published before^2^.

Participants recruited to the APPLE trial included in this biomarker discovery cohort study were characterized based on their baseline features, including demographics, disease duration, disease activity using the validated Systemic Lupus Erythematosus Disease Activity Index - SLEDAI-2K score^3^, and damage, assessed using the validated Systemic Lupus International Collaborating Clinics Damage Index- SLICC-DI^4^, in addition to baseline JSLE markers and lipid profile, as well as information about relevant JSLE treatment at baseline.

Data was collected as part of the APPLE trial protocol in a prospective manner.

***Biomarker analyses***

For proteomic analyses, we used a sensitive testing/discovery platform (Sengenics, <https://sengenics.com/i-ome-discovery-array/>) to analyse baseline serum samples. The Sengenics i-Ome Discovery platform profiles over 1,800 immune-relevant autoantibodies with high antigen specificity. A total of 579 autoantibodies identified as true signal (showing no cross-reactivity and false positive in the internal QC pooled normal group (N=6)) and were subsequently included in downstream analyses. Net intensities (NetI) were computed by subtracting local background fluorescence from foreground fluorescence for each antigen spot, measured in relative fluorescence units (RFUs). Data were analyzed under Log2 transformation and Loess normalization for consistency across array. To ensure the normalization has been performed successfully, six internal QC pooled normals were run in the experiment, and the correlation between these biological replicates were investigated. To ensure true autoantibody signal detection, non-specific binding controls were used as references. Negative correlation filter (NCF) method was applied to identify true autoantibody signal, excluding antigens highly correlated with background intensities, retaining only the least correlated.

***Data handling***

For this study, the performers/readers of the index test (the biomarker assays and subsequent computational analyses) did not have access to clinical information or reference‑standard results (CIMT progression data) at the time the index test was generated. All biomarker measurements and model development were conducted blinded to clinical outcomes to prevent bias in test interpretation. Clinical data and CIMT progression results were only linked to the index test outputs after biomarker quantification and preprocessing were completed.

The assessors of the reference standard (CIMT measurements) did not have access to index test results, as biomarker analyses were performed many years after the trial concluded. CIMT imaging and progression assessments were conducted during the original trial period, using standardised protocols, and were therefore fully blinded to any future biomarker data.

For the index test, samples with assay failure, insufficient material, or quality‑control issues were excluded from analysis and not reclassified. For the reference standard, CIMT measurements with inadequate image quality or incomplete follow‑up were treated as missing and excluded from progression‑group assignment. No indeterminate results were imputed, and all diagnostic‑accuracy analyses were performed on complete cases only.

Analyses of variability in diagnostic accuracy were conducted to assess whether the performance of the index test differed across clinically relevant subgroups. Pre‑specified analyses included evaluating diagnostic accuracy stratified by CIMT progression over 36 months in the APPLE trial. These subgroup analyses were chosen based on prior evidence suggesting potential differences in cardiovascular risk trajectories in JSLE as we published before^2^.

***Data/Statistical Analyses***

Data were assessed for normality and analyzed with parametric or nonparametric tests, as appropriate. Chi-square test was used for comparison between categorical variables. One-way ANOVA and Tukey’s range test were applied for comparisons among more than two groups. Details of statistical tests and parameters accounted for in the analyses are given in the figure legends. P < 0.05 was considered statistically significant. The autoantibody profiles of distinct CIMT progression groups identified in both the placebo and atorvastatin arms (as described and published before^2^) were compared to identify differentially expressed antigens using Empirical Bayes moderated t-test and Receiver Operating Characteristic (ROC) analysis with logistic regression model. Empirical Bayes moderated t-test borrows information across all measured features to stabilize variance estimates, thereby improving statistical power and reducing variability compared with a traditional t-test, particularly in high-dimensional datasets with relatively small sample sizes. Logistic regression was used as a predictive model to evaluate biomarker performance in distinguishing distinct JSLE groups with different CIMT progression patterns. Individual biomarkers were analyzed with univariate logistic regression, while combined markers were assessed with multivariable logistic regression through ROC analysis. Logistic regression with treatment–biomarker interaction terms were performed in the overall cohort (combined atorvastatin and placebo arms) based on 36-month change in 12 different CIMT measures. Patients were stratified into high vs. low CIMT progression groups using unsupervised hierarchical clustering of z-scored delta CIMT values. For each autoantibody, a multivariable logistic regression model was fitted with high vs. low CIMT progression as the dependent variable and baseline autoantibody level, treatment allocation (atorvastatin vs. placebo), and their interaction term (autoantibody × treatment) as independent variables. The interaction coefficient (β interaction) was used to determine whether the association between baseline autoantibody level and CIMT progression differed by treatment arm. Odds ratios (ORs) and 95% confidence intervals (CIs) were derived from model coefficients. Autoantibodies with nominal interaction P < 0.05 were considered statistically significant for differential treatment effect.

Sparse partial least squares discriminant analysis (sPLS-DA) is a supervised machine learning (ML) approach operated using the *mixOmics* package in R^5^. Model optimization was applied to select the number of components included in the sPLS-DA model. The models with optimized component number that gave the lowest overall estimation error rate were selected as optimal, giving the best discriminatory performance for further analysis. The separation between groups of samples was presented by projecting each sample into the subspace constructed of component 1 and component 2. The top-weighted parameters in each component were selected and presented by variable loading plots.

Diagnostic accuracy of the index test was evaluated using receiver operating characteristic (ROC) curve analysis. ROC curves were generated by plotting sensitivity against 1–specificity across all possible biomarker thresholds. The area under the ROC curve (AUC) was used as the primary measure of discriminative ability, with higher AUC values indicating better performance in distinguishing high versus low CIMT progression. AUC of each model were computed with the *pROC* package in R^6^. Youden method was applied to determine the optimal balance between sensitivity and specificity, providing the optimal model accuracy of the model. To enable integrated analysis relevant for CVD-risk estimation, lipid metabolomics and autoantibody profiles identified in matched baseline serum samples, together with disease parameters at baseline were incorporated into the sPLS-DA prediction models.

***Model validation***

Five-fold cross-validation was applied for the sPLS-DA models with the *caret* package in R^7^. Data were randomly partitioned into 5 groups of almost equal size. Four groups were used as training data for model construction and the remaining group was used as validation data. The process was repeated for all 5 folds until each observation in the data was used for validation purpose once. The average performance of the 5 models was used as the result of the 5-fold cross-validation.

***Protein-protein interaction (PPI) analysis***

PPI enrichment analysis was performed in Metascape^8^ for each protein/autoantigen list using a combination of curated databases, including STRING, BioGRID, OmniPath, and InWeb_IM. To ensure relevance to direct physical interactions, only experimentally validated physical interactions were kept from STRING (interaction score > 0.132) and BioGRID. Because of the low number of targets, we could not perform PPI enrichment analyses in the placebo arm cohort.

***Functional annotations***

The functional information for the identified autoantigens was obtained from the UniProt and KEGG databases^9,10^.

***Exploratory correlation analysis***

Exploratory correlations between baseline serum autoantibody signatures and clinical features in the APPLE trial sub-cohort (N = 94) were assessed using Spearman’s rank correlation for the placebo and atorvastatin arms, respectively. Clinical variables included age, sex, disease duration, body mass index (BMI), SLEDAI-2k, SLICC, puberty status, low-density lipoprotein and total cholesterol, inflammatory markers (high-sensitivity C-reactive protein and erythrocyte sedimentation rate), complement levels (C3 and C4), red blood cell count, white blood cell count, platelet count, serological status (anti-dsDNA, anti-Ro and anti-La antibodies), proteinuria, and baseline corticosteroid use. correlations meeting nominal significance thresholds (P < 0.05 and |ρ| > 0.30) were visualized using chord plots generated with the *circlize* R package^11^. No adjustment for multiple testing was applied due to the exploratory nature of the analysis.

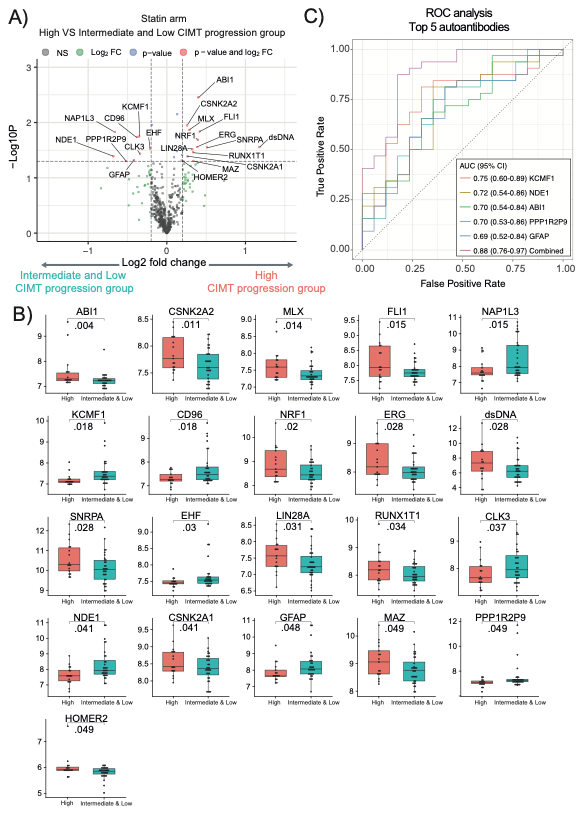

**eFigure 1.** Baseline serum autoantibodies (N=579 after data cleaning) comparisons between distinct CIMT progression groups (high vs. low and intermediate, identifying non-responders vs. responders and partial responders to statin) - atorvastatin arm.

*Legend:* A) Volcano plot displaying fold change of all autoantibodies and Log10 p values comparing high (N=17) vs. low and intermediate combined (N=32) CIMT progression groups - atorvastatin arm (*P* < 0.05; log2(fold change)>0.2). B) Box and whisker plots showing the significantly different autoantibody levels of the high vs. low CIMT progression groups - atorvastatin arm. Empirical Bayes moderated t-test. C) Combined and separate ROC analyses for discriminating high vs. low CIMT progression groups, using the top five autoantibodies (ranked by individual AUC, all with AUC>69%). *Abbreviations*: AUC - area under the curve; CI- 95% confidence interval; CIMT- carotid intima-media thickness; ROC- Receiver Operating Characteristic.

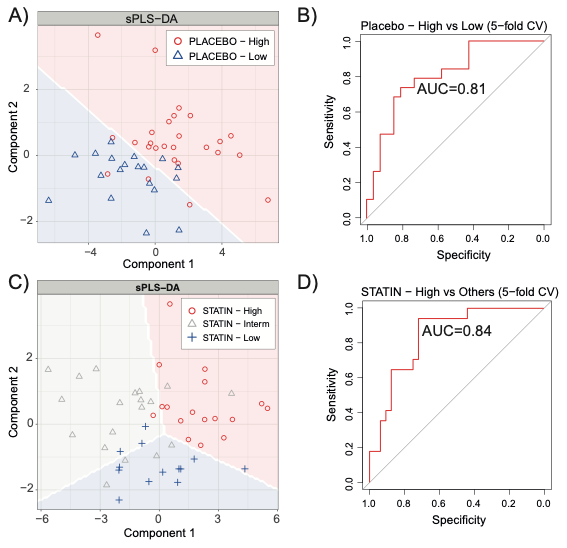

**eFigure 2.** Sparse Partial Least Squares Discriminant Analysis (sPLS-DA) using identified autoantibody signature, identified lipid metabolomic signature and clinical variables to predict CVD-risk in placebo and statin arms.

*Legend:* **A)** sPLS-DA plot distinguishing individuals with high vs. low CIMT progression group in the placebo arm. Individual distribution points with predictive backgrounds for the high and low CIMT progression group are plotted in red and blue, respectively. **B)** ROC curve showing the performance of placebo combined signature model predicting high CIMT progression group in the placebo arm under 5-fold cross-validation. **C)** sPLS-DA plot distinguishing individuals with high, intermediate, and low CIMT progression in the statin arm. Individual distribution points with predictive background for the high, intermediate, and low CIMT progression group are plotted in red, grey, and blue, respectively. **D)** ROC curve showing the performance of atorvastatin combined signature model predicting high CIMT progression group in the atorvastatin arm under 5-fold cross-validation.

*Abbreviations:* AUC - area under the curve; CIMT- carotid intima-media thickness; ROC- Receiver Operating Characteristic.

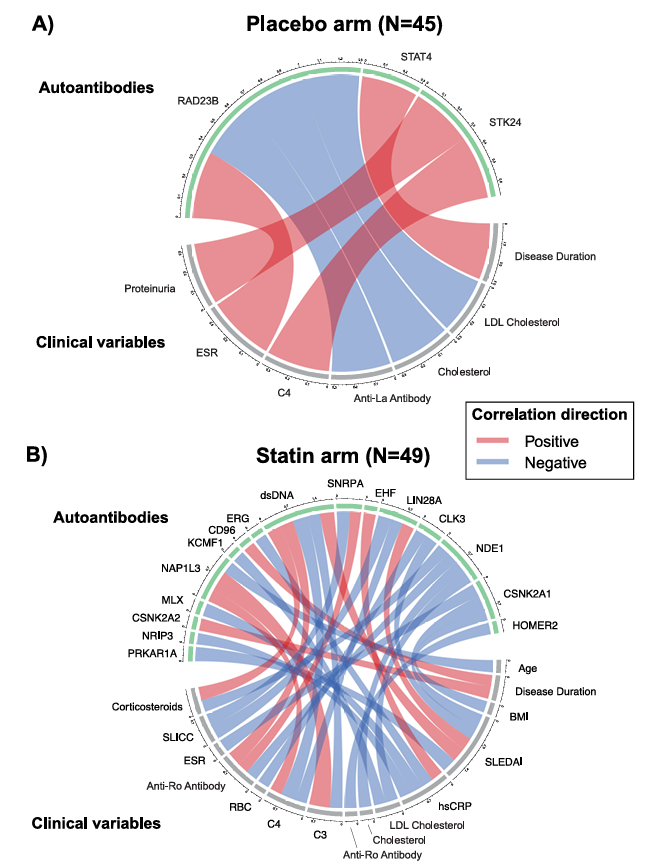

**eFigure 3.** Chord plot showing exploratory correlations between baseline autoantibody signature and clinical features in the APPLE trial sub-cohort (N=94)

*Legend:* A) Correlations between baseline serum autoantibodies in the placebo atherosclerosis progression signature and various JSLE-related clinical variables at baseline (Placebo arm, N=45). B) Correlations between baseline serum autoantibodies in the atorvastatin response signature and the same JSLE-related clinical variables at baseline (atorvastatin arm, N=49). Spearman’s rank correlation coefficients are represented as connecting lines between the autoantibody section and the clinical variables section. Only correlations with P < 0.05 and |Rho|>0.3 are shown (*P* values were not adjusted due to the exploratory nature of the analysis). Red line represents a positive correlation and blue line a negative correlation. The width of the connecting lines indicates the strength of the correlation.

*Abbreviations*: BMI – body mass index; C3 – complement component 3; C4 – complement component 4; ESR – erythrocyte sedimentation rate; hsCRP – high-sensitivity C-reactive protein; LDL – low-density lipoprotein; RBC – red blood cell count; SLEDAI – Systemic Lupus Erythematosus Disease Activity Index; SLICC/ACR-DI – Systemic Lupus International Collaborating Clinics/American College of Rheumatology Damage Index.

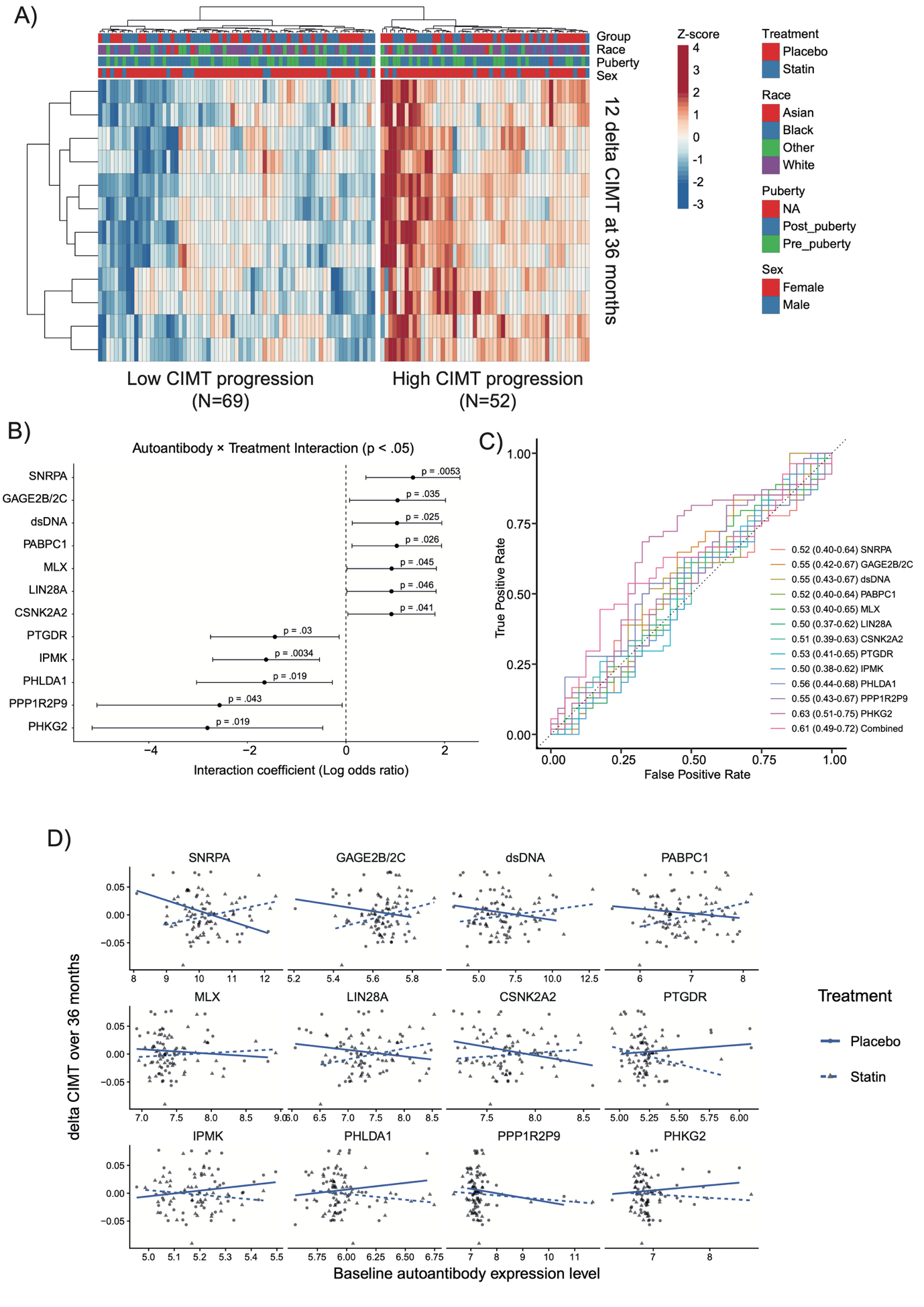

1. **eFigure 4.** Treatment–biomarker interaction logistic regression analysis in the APPLE trial sub-cohort.

*Legend:* **A)** JSLE stratification by delta CIMT (12 measurements) at baseline vs 36 months in the APPLE subcohort (placebo and atorvastatin arms, N=121). Heatmap displaying delta CIMT (z scored) from JSLE patients stratified by unsupervised hierarchical clustering. Unsupervised hierarchical clustering identified distinct high (N=52) and low (N=69) CIMT progression groups. Each column represents an individual JSLE patient. **B)** Forest plot showing the interaction coefficient (β_interaction, log odds ratio) and 95% confidence intervals from logistic regression models assessing the association between baseline autoantibody levels and CIMT progression (high vs. low), including treatment group (atorvastatin vs. placebo, N=94) and their interaction term. Only the top 12 autoantibodies with nominal interaction (P < 0.05) in logistic regression models were shown. **C)** Combined and separate ROC analyses for discriminating high vs. low CIMT progression groups, using the top 12 autoantibodies with nominal interaction (P < 0.05) in logistic regression model. **D)** Scatter plots with fitted lines illustrating the relationship between baseline autoantibody levels and change in mean–mean common carotid intima–media thickness (primary end point of the APPLE trial) over 36 months. Panels correspond to the top 12 autoantibodies with nominal interaction (P < 0.05) in logistic regression model, stratified by treatment group (atorvastatin vs. placebo).

*Abbreviations:* APPLE, Atherosclerosis Prevention in Paediatric Lupus Erythematosus; AUC - area under the curve; β_interaction, interaction coefficient; CI- 95% confidence interval; CIMT, carotid intima–media thickness; delta CIMT, change in carotid intima–media thickness over 36 months; JSLE, juvenile-onset systemic lupus erythematosus; ROC- Receiver Operating Characteristic.

**e****Table 1** Names and functions of the top significantly differentially expressed autoantibodies identified in the high vs. low CIMT progression groups in the placebo arm.

| Name | Full Name | Function |
| --- | --- | --- |
| **RAD23B** | RAD23 homolog B | Involved in the nucleotide excision repair (NER). |
| **HDAC4** | Histone Deacetylase 4 | Role in altering chromosome structure and affects transcription factor access to DNA. Contributes to angiogenesis. |
| **STAT4** | Signal Transducer and Activator of Transcription 4 | Response to cytokines and growth factors. Essential for mediating responses to IL12 in lymphocytes and regulating the differentiation of T helper cells. STAT4 polymorphism increase the risk of SLE ^11^. STAT4-dependnet neutrophil activation has been found to have pro-atherogenic role in mice ^12^. |
| **SEPTIN9** | Septin 9 | Involved in cytokinesis and cell cycle control. Also implicated in cancer development, particularly in methylating processes associated with colorectal cancer. |
| **STK24** | Serine/Threonine kinase 24 | Involved in upstream of mitogen-activated protein kinase (MAPK) signalling. Functions in the regulation of apoptosis and cell proliferation, and it is involved in stress responses and cell cycle control. |
| **NFIA** | Nuclear Factor I A | Recognizes and binds the palindromic sequence 5'-TTGGCNNNNNGCCAA-3' present in viral and cellular promoters and in the origin of replication of adenovirus type 2. These proteins are individually capable of activating transcription and replication. |

*Legend:* Functional information was obtained from UniProt and KEGG databases^9,10^. Autoantibodies with significantly elevated levels (P<0.05; log2(fold change)>0.2) in the high CIMT progression group are highlighted in Red, and the ones with significantly decreased levels (P<0.05; log2(fold change)>0.2) are in Black.

**eTable 2.** Performance of the top significantly differentially expressed autoantibodies in stratifying atherosclerosis progression risk in the placebo arm.

| Markers | Cut-off | Number of patients with high atherosclerosis progression over 3 years (N=26) stratified in the high-risk group | Number patients with low atherosclerosis progression over 3 years (N=19) stratified as low-risk group | Sensitivity | Specificity | AUC (95% CI) |
| --- | --- | --- | --- | --- | --- | --- |
| RAD23B | 7.1 | 23 | 10 | 0.88 | 0.53 | 0.69  (0.51-0.83) |
| HDAC4 | 7.41 | 18 | 14 | 0.69 | 0.74 | 0.69  (0.52-0.82) |
| STAT4 | 8.07 | 18 | 13 | 0.69 | 0.68 | 0.66  (0.48-0.80) |
| SEPTIN9 | 6.56 | 12 | 17 | 0.46 | 0.89 | 0.65  (0.46-0.79) |
| STK24 | 7.21 | 9 | 18 | 0.35 | 0.95 | 0.61  (0.43-0.76) |
| NFIA | 7.43 | 9 | 15 | 0.35 | 0.79 | 0.56  (0.40-0.74) |

*Legend:* Summary of the performance metrics for six autoantibodies (RAD23B, HDAC4, STAT4, SEPTIN9, STK24, NFIA) differentially expressed in the high (N=26) vs. low (N=19) atherosclerosis progression groups in the placebo arm. The AUC were calculated using logistic regression ROC analysis. The optimal sensitivity, specificity and cut-off values were determined using Youden index method. Autoantibodies with significantly elevated levels (P<0.05; log2(fold change)>0.2) in the high CIMT progression group are highlighted in Red, and the ones with significantly decreased levels (P<0.05; log2(fold change)>0.2) are in Black.

**eTable 3.** Names and functions of the top significantly differentially expressed autoantibodies in stratifying atherosclerosis progression risk in the atorvastatin arm (includes both comparisons between the high vs. low CIMT progression groups (non-responders vs. responders to statins) and the combined high vs. low/moderate progression groups (non-responders vs. responders and partial responders to statin).

|  | Name | Full Name | Function |
| --- | --- | --- | --- |
| **1** | **PRKAR1A** | cAMP-dependent protein kinase type I-alpha regulatory subunit | Regulatory subunit of the cAMP-dependent protein kinases involved in cAMP signaling in cells |
| **2** | **ATP5B** | ATP synthase subunit beta | Role in ATP synthesis in mitochondria |
| **3** | **ABI1** | Abelson interactor 1 | Negative regulation of cell growth and transformation. Loss-of-function of MBNL1 activates vascular smooth muscle cells (VSMC) to macrophage-like cells transformation to promote atherogenesis through regulating Abi1 RNA splicing ^13^. |
| **4** | **PDK4** | Pyruvate Dehydrogenase kinase isozyme 4 | Role in regulation of glucose and fatty acid metabolism and homeostasis. PDK4 contributes vascular calcification by disrupting with autophagic activity and metabolic reprogramming ^14^. |
| **5** | **CSNK2A2** | Casein kinase II subunit alpha' | Involved in cell cycle regulation, DNA repair, and apoptosis. CK2 inhibition prevents the accumulation of vascular smooth muscle cells (VSMC) within the neointimal compartment, a cause of accelerated atherosclerosis ^15^. Associated with CVD and identified as potential therapeutic target ^16^. |
| **6** | **NRIP3** | Nuclear receptor interacting protein 3 | Unclear |
| **7** | **NDE1** | Nuclear distribution protein nudE homolog 1 | Required for centrosome duplication and formation and function of the mitotic spindle. Essential for the development of the cerebral cortex. |
| **8** | **KCMF1** | Potassium channel modulatory factor 1 | Enables ubiquitin protein ligase activity. |
| **9** | **PPP1R2P9** | Protein phosphatase 1 regulatory subunit 2 pseudogene 9 | Regulates protein phosphatase 1. |
| **10** | **LIN28A** | Lin-28 homolog A | RNA-binding protein which inhibits processing of pre-let-7 miRNAs and regulates translation of mRNAs that control developmental timing, pluripotency, and metabolism. Lin28A up‐regulation is linked to atherosclerosis lesions restenosis ^17^. |
| **11** | **MLX** | Max-like protein X | Transcription factor. Plays a role in transcriptional activation of glycolytic target genes. Involved in glucose-responsive gene regulation. |
| **12** | **GFAP** | Glial fibrillary acidic protein | Cell-specific marker that, during the development of the central nervous system, distinguishes astrocytes from other glial cells. |
| **13** | **NAP1L3** | Nucleosome assembly protein 1-like 3 | Involved in chromatin assembly and gene regulation. |
| **14** | **EHF** | ETS homologous factor | Regulates epithelial cell differentiation and proliferation. |
| **15** | **dsDNA** | Double-stranded DNA | Anti-dsDNA antibodies are highly specific markers of lupus. Known effect on increased CVD risk in lupus ^18^. |
| **16** | **FLI1** | Friend leukemia integration 1 transcription factor | Sequence-specific transcriptional activator. |
| **17** | **CD96** | T-cell surface protein tactile | Role in T-cell activation. Typically expressed on NK cells and T cells |
| **18** | **CLK3** | CDC-like kinase 3 | Kinase involved in regulating pre-mRNA splicing. |
| **19** | **HOMER2** | Homer protein homolog 2 | Postsynaptic density scaffolding protein. Aids the coupling of surface receptors to intracellular calcium release. Negatively regulates T cell activation. |
| **20** | **SNRPA** | U1 small nuclear ribonucleoprotein A | Essential for RNA splicing and spliceosome assembly. |
| **21** | **ERG** | ETS-related gene | Transcriptional regulator essential for endothelial homeostasis. ERG inhibits vascular inflammation by repressing NF-kappaB activation and proinflammatory gene expression in endothelial cells. Loss of ERG expression is associated with diseases including atherosclerosis ^19^. |
| **22** | **RUNX1T1** | RUNX1 translocation partner 1 | Transcriptional corepressor that regulates gene expression. Role in differentiation and proliferation of adipocytes. Potential therapeutic target for cardiovascular disease ^20^. |
| **23** | **MAZ** | MYC-associated zinc finger protein | Transcriptional regulator involved in transcription initiation and termination. MAZ-Ab identified as potential marker of atherosclerosis ^21^. |
| **24** | **CSNK2A1** | Casein kinase II subunit alpha | Kinase that phosphorylates acidic proteins. Involved in cell cycle control, apoptosis, and circadian rhythms. CK2 inhibition prevents the accumulation of vascular smooth muscle cells (VSMC) within the neointimal compartment, a cause of accelerated atherosclerosis ^15^. Associated with CVD and identified as potential therapeutic target ^16^. |
| **25** | **NRF1** | Nuclear respiratory factor 1 | Regulates mitochondrial function and energy metabolism. |
| **26** | **BATF** | Basic Leucine Zipper ATF-Like Transcription Factor | Thought to be a negative regulator of AP-1/ATF transcriptional events. |
| **27** | **NUDT2** | Nudix Hydrolase 2 | Novel complex formation enhancing factor regulating mTORC1-Rag GTPase signaling that is crucial for cell growth control |

*Legend:* Functional information was obtained from UniProt and KEGG databases ^9,10^. Autoantibodies with significantly elevated levels (P<0.05; log2(fold change)>0.2) in the high CIMT progression group are highlighted in Red, and the ones with significantly decreased levels (P<0.05; log2(fold change)>0.2) are in Black.

**eTable 4.** Performance of the top significantly differentially expressed autoantibodies in stratifying atherosclerosis progression risk in the atorvastatin arm (includes both comparisons between the high vs. low CIMT progression groups (non-responders vs. responders to statins), and the combined high vs. low/moderate progression groups (non-responders vs. responders and partial responders to statin).

| Markers | Cut-off | Number of patients who progressed despite statins classified as non-responders by the test | Number of patients responsive to statins classified as responders by the test | Sensitivity | Specificity | AUC  (95% CI) |
| --- | --- | --- | --- | --- | --- | --- |
| Non-responders (high atherosclerosis progression) (N=17) vs. Responders (low atherosclerosis progression) to statin (N=13) | | | | | | |
| PRKAR1A | 7.34 | 13 | 11 | 0.76 | 0.85 | 0.74  (0.53-0.90) |
| ABI1 | 7.27 | 11 | 11 | 0.65 | 0.85 | 0.74  (0.52-0.89) |
| NRIP3 | 6.11 | 7 | 13 | 0.41 | 1 | 0.71  (0.50-0.86) |
| CSNK2A2 | 7.59 | 13 | 7 | 0.76 | 0.54 | 0.67  (0.45-0.83) |
| PDK4 | 7.24 | 9 | 10 | 0.53 | 0.77 | 0.62  (0.40-0.79) |
| ATP5B | 7.19 | 10 | 9 | 0.59 | 0.69 | 0.60  (0.38-0.78) |
| BATF | 7.22 | 10 | 9 | 0.59 | 0.69 | 0.59  (0.37-0.77) |
| NUDT2 | 6.95 | 9 | 8 | 0.53 | 0.62 | 0.47  (0.26-0.68) |
| Non-responders (high atherosclerosis progression) (N=17) vs. Responders and Partial Responders to statin (low atherosclerosis progression and moderate atherosclerosis progression) (N=32) | | | | | | |
| KCMF1 | 7.19 | 12 | 26 | 0.71 | 0.81 | 0.75  (0.60-0.89) |
| NDE1 | 7.64 | 11 | 26 | 0.65 | 0.81 | 0.72  (0.54-0.86) |
| ABI1 | 7.26 | 12 | 21 | 0.71 | 0.66 | 0.7  (0.54-0.84) |
| PPP1R2P9 | 7.09 | 10 | 26 | 0.59 | 0.81 | 0.7  (0.53-0.86) |
| GFAP | 7.73 | 10 | 26 | 0.59 | 0.81 | 0.69  (0.52-0.84) |
| LIN28A | 7.25 | 13 | 18 | 0.76 | 0.56 | 0.68  (0.51-0.84) |
| NAP1L3 | 7.77 | 12 | 19 | 0.71 | 0.59 | 0.68  (0.51-0.82) |
| EHF | 7.53 | 14 | 18 | 0.82 | 0.56 | 0.68  (0.52-0.83) |
| HOMER2 | 5.86 | 15 | 17 | 0.88 | 0.53 | 0.68  (0.52-0.84) |
| CSNK2A2 | 7.59 | 13 | 16 | 0.76 | 0.5 | 0.67  (0.51-0.82) |
| CD96 | 7.77 | 17 | 9 | 1 | 0.28 | 0.67  (0.50-0.82) |
| SNRPA | 9.61 | 17 | 9 | 1 | 0.28 | 0.67  (0.50-0.82) |
| MLX | 7.56 | 9 | 25 | 0.53 | 0.78 | 0.66  (0.50-0.83) |
| dsDNA | 7.24 | 9 | 26 | 0.53 | 0.81 | 0.65  (0.53-0.82) |
| FLI1 | 8.33 | 7 | 30 | 0.41 | 0.94 | 0.65  (0.54-0.82) |
| CLK3 | 7.7 | 10 | 23 | 0.59 | 0.72 | 0.65  (0.51-0.79) |
| MAZ | 9.03 | 9 | 24 | 0.53 | 0.75 | 0.64  (0.53-0.80) |
| NRF1 | 9.13 | 6 | 30 | 0.35 | 0.94 | 0.64  (0.55-0.81) |
| ERG | 8.72 | 6 | 30 | 0.35 | 0.94 | 0.63  (0.55-0.80) |
| RUNX1T1 | 8.07 | 11 | 20 | 0.65 | 0.62 | 0.62  (0.56-0.79) |
| CSNK2A1 | 8.83 | 6 | 29 | 0.35 | 0.91 | 0.62  (0.54-0.79) |

*Legend:* Summary of the performance metrics of the 29 novel autoantibodies identified in the atorvastatin arm, by comparing the high (N=17) vs. low (N=13) atherosclerosis progression groups (8 antibodies) or by comparing the high (N=17) vs. low and intermediate/moderate (N=32) atherosclerosis progression groups (21 antibodies, with two overlapping between the two signatures). The AUC were calculated using logistics regression ROC analysis. The optimal sensitivity, specificity and cut-off values were determined using Youden index method. Autoantibodies with significantly elevated levels in high CIMT progression group are highlighted in Red and the ones significantly decreased are in Black.

**eTable 5.** Names and functions of the top autoantibodies (P < 0.05) identified in treatment–biomarker interaction logistic regression analysis.

| Name | Full Name | Function |
| --- | --- | --- |
| **SNRPA** | U1 small nuclear ribonucleoprotein A | Essential for RNA splicing and spliceosome assembly. |
| **GAGE2B/2C** | G antigen 2B/ 2C | Recognized on melanomas by autologous cytolytic T-lymphocytes. |
| **dsDNA** | Double-stranded DNA | Anti-dsDNA antibodies are highly specific markers of lupus. Known effect on increased CVD risk in lupus ^18^. |
| **PABPC1** | Poly(A)-binding protein cytoplasmic 1 | Binds polyadenylated mRNA; involved in regulation of mRNA stability and translation initiation. |
| **MLX** | Max-like protein X | Transcription factor. Plays a role in transcriptional activation of glycolytic target genes. Involved in glucose-responsive gene regulation. |
| **LIN28A** | Lin-28 homolog A | RNA-binding protein which inhibits processing of pre-let-7 miRNAs and regulates translation of mRNAs that control developmental timing, pluripotency, and metabolism. Lin28A up‐regulation is linked to atherosclerosis lesions restenosis ^17^. |
| **CSNK2A2** | Casein kinase II subunit alpha' | Involved in cell cycle regulation, DNA repair, and apoptosis. CK2 inhibition prevents the accumulation of vascular smooth muscle cells (VSMC) within the neointimal compartment, a cause of accelerated atherosclerosis ^15^. Associated with CVD and identified as potential therapeutic target ^16^. |
| **PTGDR** | Prostaglandin D2 receptor | G protein-coupled prostanoid receptor binding prostaglandin D₂; mediates PGD₂ signaling via cAMP modulation in immune and vascular cells. |
| **IPMK** | Inositol polyphosphate multikinase | Inositol phosphate kinase that phosphorylates InsP₃/InsP₄ to higher inositol phosphates (e.g., InsP₅); plays roles in inositol phosphate biosynthesis and nuclear signaling. |
| **PHLDA1** | Pleckstrin homology-like domain family A member 1 | Involved in regulating apoptosis, cell proliferation, and stress responses. |
| **PPP1R2P9** | Protein phosphatase 1 regulatory subunit 2 pseudogene 9 | Regulates protein phosphatase 1. |
| **PHKG2** | Phosphorylase kinase catalytic subunit gamma 2 | Catalytic gamma subunit of phosphorylase kinase, regulating glycogen breakdown (glycogenolysis) through phosphorylation of glycogen phosphorylase. |

*Legend:* Functional information was obtained from UniProt and KEGG databases ^9,10^. Autoantibodies overlapping with those detected in the separate within‑arm analyses are highlighted in Blue.
